# The analysis of heart rate variability and accelerometer mobility data in the assessment of symptom severity in psychotic disorder patients using a wearable Polar H10 sensor

**DOI:** 10.1101/2023.08.04.23293640

**Authors:** Kamil Książek, Wilhelm Masarczyk, Przemysław Głomb, Michał Romaszewski, Iga Stokłosa, Piotr Ścisło, Paweł Dębski, Robert Pudlo, Krisztián Buza, Piotr Gorczyca, Magdalena Piegza

## Abstract

**Background and Objective:** Advancement in mental health care requires easily accessible diagnostic and treatment assessment tools. There is an ongoing search for biomarkers that would enable objectification and automatization of the diagnostic and treatment process dependent on a psychiatric interview. Current wearable technology and computational methods make it possible to incorporate heart rate variability (HRV), an indicator of autonomic nervous system functioning and a potential biomarker of disease severity in mental disorders, into accessible diagnostic and treatment assessment frameworks.

**Methods:** We used a commercially available electrocardiography (ECG) chest strap with a built-in accelerometer, i.e. Polar H10, to record R-R intervals and activity of 30 hospitalized schizophrenia or bipolar disorder patients and 30 control participants for 1.5-2 hours time periods. We performed an analysis to assess the relationship between HRV and the Positive and Negative Syndrome Scale (PANSS) test scores. The source code for the reproduction of all experiments is available on GitHub while the dataset is available in Zenodo.

**Results and Conclusions:** Mean HRV values were lower in the patient group and negatively correlated with the results of the PANSS general subcategory. For the control group, we also discovered the inversely proportional dependency between the mobility coefficient based on accelerometer data and HRV. This relationship was less pronounced for the patient group. This indicates that HRV and mobility may be promising markers in disease diagnosis.

## 1 Introduction

Mental disorders are among the crucial global health problems affecting individuals’ cognition, behavior and emotional regulation. According to the World Health Organization (WHO), 1 in 8 people in the world live with a mental disorder and most have no access to effective care [World Health Organization, 2022]. Schizophrenia and bipolar disorder constitute a major part of the problem due to their chronic, progressive nature and highly negative impact on quality of life [Evans et al., 2007]. Their prevalence in Western populations is estimated at 1%.

Schizophrenia is characterized by global information processing disintegration in the central nervous system (CNS) that clinically manifests itself in the dimension of positive and negative symptoms and cognitive deficits [Kahn et al., 2015]. Positive symptoms include delusions, thought disturbances, and hallucinations. Negative symptoms include emotional rigidity, difficulty in establishing emotional contact, speech poverty, and withdrawal from social contacts. Cognitive deficits include impaired attention, memory, learning skills, and a general decline in intellectual functioning [McCutcheon et al., 2020].

Bipolar disorder (BD) shares several characteristics with schizophrenia, including the occurrence of psychotic episodes. BD can also be considered in terms of the dimensions of positive and negative symptoms but the core feature of the disease is the presence of manic or depressive mood disturbances.

The most common reason for admission of BD or schizophrenia patients to a psychiatric ward is a severe deterioration in the positive symptoms and everyday functioning [Olfson et al., 2011]. The diagnostic criteria for both schizophrenia and BD are defined by the International Classification of Diseases (ICD-11) and the Diagnostic and Statistical Manual of Mental Disorders (DSM-5). Although the diagnostics criteria are well established, the occurrence of disease symptoms is observed subjectively by a specialist, who must refrain himself from the influence of his mood, expectations, and preconceptions about the patient. These factors may have an unknown, highly unwanted impact on the diagnosis and treatment. Currently, there are no well-established objective and observer-independent diagnostic tools for schizophrenia or any other mental disorder. Additionally, while diagnostics criteria are clearly defined, there is a surprising lack of well-defined criteria for remission and disease severity [Lambert et al., 2010]. The evaluation is influenced by many factors: the quality of the therapeutic contact, the patient’s willingness to cooperate, the amount of time the specialist can spend with the patient, the timing of the assessment, and also the experience and mood state of the specialist.

There are tools that support the evaluation of the disease severity, such as the Positive and Negative Syndrome Scale (PANSS) [Kay et al., 1987]. Its results are, to a certain point, subjective, as it requires a trained physician to rate symptoms based on a structured interview and answer sheet. The method is personnel and time-consuming but can evaluate a change in symptom severity in consecutive days [Opler et al., 2017]. In addition, structured interviews can give different results than semi-structured open interviews. There is a difference in understanding of the questions and answers because a structured interview does not account for different circumstances surrounding a patient, as well as a phenomenologically oriented conversational semi-structured interview [Nordgaard et al., 2013].

In order to provide objective, easily accessible information about the intensity of the disease and its changes, there are attempts to use biomarkers in psychiatry for prediction, diagnostics, and treatment monitoring. This often requires access to complex diagnostic tools such as magnetic resonance imaging (MRI) or genetic sequencing and can be costly and time-consuming with very little to no use in a clinical setting [García-Gutiérrez et al., 2020].

There are initial attempts in automated ML-assisted monitoring in psychiatry that could provide objective, easy-to-obtain information about the current severity of symptoms and the change in severity in short timespans under treatment, but there is still room for improvement [Chekroud et al., 2021].

Heart rate variability (HRV) is a promising potential biomarker of disease severity in schizophrenia and BD that is obtainable through wearable electrocardiogram monitors and, as a data source, is well suited for ML processing [Benjamin et al., 2021].

HRV is a derivative function of the concurring sympathetic and parasympathetic networks of the autonomic nervous system. Autonomic dysfunction plays an important part in psychiatric diseases, including schizophrenia. This can be potentially explained by the Neurovisceral Integration Hypothesis, in which the prefrontal cortex (PFC) exerts tonic inhibition over limbic brain regions that typically suppress parasympathetic responses and activate sympathetic responses. In schizophrenia, deficits in the PFC may lead to disinhibition of the amygdala and medullary regions during emotional processing. This may potentially result in suppression of parasympathetic activity and stimulation of sympathetic activity, causing increased heart rate and decreased heart rate variability [Stogios et al., 2021]. Depending on the HRV analysis modality, it is possible to extract information that dominantly represents the parasympathetic part of the autonomic activity (i.e. root mean square of successive variances of N-N intervals, high-frequency power) or a mix of sympathetic and parasympathetic activity (i.e. standard deviation of N-N intervals, low-frequency power) [Matusik et al., 2023].

However, there are practical issues with HRV acquisition. The electrocardiogram (ECG) signal is prone to artifacts due to the movement of the electrodes and body. Also, additional ectopic beats and arrhythmia, in general, can make it difficult or even impossible to acquire data that can be interpreted as a derivative of autonomic activity because the intrinsic electrical activity of the heart becomes the leading source of pacing, effectively hiding the influence of the autonomic nervous system (ANS) [Olshansky, 2018].

To avoid movement artifacts during signal acquisition, it is important to use ECG devices of sufficient quality. The consumer-grade Polar H10 chest strap tends to outclass other consumer and medical-grade devices, especially during body movement, also maintaining an acceptable price/quality ratio for its common use in an outpatient setting [Gilgen-Ammann et al., 2019, Speer et al., 2020, Hinde et al., 2021]. In addition, the device has a built-in accelerometer, the data of which has the potential to help in psychiatric assessment [Reinertsen et al., 2017].

In this paper, we focused on supporting objectification and automatization of the psychotic disorder diagnostic with HRV and accelerometer activity as biomarkers. We analyzed the relationships between these factors and PANSS test scores and compared them between the control and treatment groups. Our main contributions are as follows:

1. We prepared a dataset consisting of measurements from 30 schizophrenia and bipolar disorder patients and a control group of 30 people. For each person, the data was collected from more than 1-hour readings using Polar H10 wearable devices. To make the results fully reproducible, we published the dataset Książek et al. [2023b] and experimental code Książek et al. [2023a] under an open license.
2. We performed a statistical analysis of the dataset, including the study of relationships between HRV values and disease severity in terms of the PANSS (Positive and Negative Syndrome Scale [Kay et al., 1987]) as well as differences between patients and control group.
3. We investigated the relationship between HRV values and accelerometer-measured mobility. By using a period of measurements of more than one hour with a suggested movement pattern (see Section 2) we registered a large amount of data that contained both stationary and non-stationary episodes. This allowed us to investigate the dynamics of HRV values within each patient’s ability to move or rest, which may be caused by the severity of the psychotic disorder. We discuss dependencies between patients’ activity and HRV as well as differences in patterns between people from the treatment and control groups. The results indicate that the relationship between patients’ HRV and mobility can significantly differ from that observed in the general population [Kazmi et al., 2016, Yiiong et al., 2019].

### 1.1 Related work

#### 1.1.1 Background

There exist multiple methods of HRV calculation [Bravi et al., 2011, Quintana et al., 2016]. One of the standard approaches divides the time series into multiple parts and applies spectrum analysis methods, e.g. the Fast Fourier Transform. Another class of methods is related to time domain analysis [Bravi et al., 2011], including the calculation of the RMSSD, i.e. the Root Mean Square of the Differences between consecutive normal (non-anomalous) R-R intervals, called N-N intervals. Following the recommendation of the [Task Force of the European Society of Cardiology and the North American Society of Pacing and Electrophysiology, 1996], frequency domain methods should be used for short measurements, e.g. 5 minutes. The time domain approach is recommended for long (e.g. nightly) measurements. However, time domain methods can also be applied for short-term recordings but the results are harder to interpret from the physiological point of view.

#### 1.1.2 Other papers analyzing HRV

HRV analysis is an active area of research. [Benjamin et al., 2021] describes a study that most influenced our work, with 236 participants, including 35 patients with schizophrenia, 52 with bipolar disorder and 149 healthy people. 5-minute measurement periods are used. Results indicate that HRV values are correlated with PANSS, the Global Assessment of Functioning (GAF), and the Young Mania Rating Scale (YMRS). In [Bengtsson et al., 2021], the authors describe a study with 37 schizophrenia patients, 43 patients with depression and 64 healthy people. They attempt to assess the influence of anticholinergic drugs and physical activity on HRV values. Accelerometer is used to assess the presence of physical activity and compare patterns between groups. [Quintana et al., 2016] provides a guide for measuring HRV in psychiatry, including instructions for data collection, artifact correction, and HRV calculation. The authors emphasize general requirements for results reproducibility. [Henry et al., 2010] compares the HRV values of 23 patients with bipolar disorder and 14 patients with schizophrenia. Several ways of HRV calculation are analyzed, including time and frequency domain and nonlinear methods. Also, in this case, 5-minute measurement periods are used. Results are focused on correlations between HRV, YRMS, and the Brief Psychiatric Rating Scale (BPRS). PANSS scores were not considered in this study. The work [Chang et al., 2009] contains a study on schizophrenia patients and HRV patterns with 30 patients and 30 healthy people. Results include multivariate analysis of variance between groups and comparison of HRV metrics. As in the previous work, 5-minute measurement periods are used. The authors of [Clamor et al., 2016] perform a meta-analysis of 34 papers related to the analysis of HRV for patients with schizophrenia. They compare differences between the results of particular studies and claim that patients with schizophrenia have lower HRV values than the control groups.

## 2 Methods

In this study, we recruited 30 adult patients with diagnosed schizophrenia or bipolar disorder hospitalized in the Psychiatric Department in Tarnowskie Góry. The control group consisted of 30 adults without any current psychiatric condition with similar age and sex structure. The minimal age in the treatment and the control were 20 and 24, respectively, while the maximal age was 69 in both groups. The demographic structure in the tested groups is presented in Table 1 while the distribution of age with kernel density estimation curves is shown in Figure 1.

Participants were informed and asked for written consent prior to admission to the experiment. The experimental procedure in the present study received approval from the Bioethics Committee of the Medical University of Silesia, No.: BNW/NWN/0052/KB1/135/I/22/23.

**Table 1:** The demographic structure of the study.

| Sex structure (women/men) |  |
| --- | --- |
| treatment | 16 / 14 |
| control | 17 / 13 |
| Age structure (mean ± std. dev.) | |
| treatment | 41.43 ± 13.06 |
| control | 43.20 ± 14.06 |
| Disease structure |  |
| schizophrenia | 23 |
| bipolar disorder | 7 |
| Additional diagnosis in the treatment group (women/men) |  |
| drugs and other psychoactive substances | 0 / 7 |
| hypnotics/sedatives | 0 / 1 |

**Figure 1:**
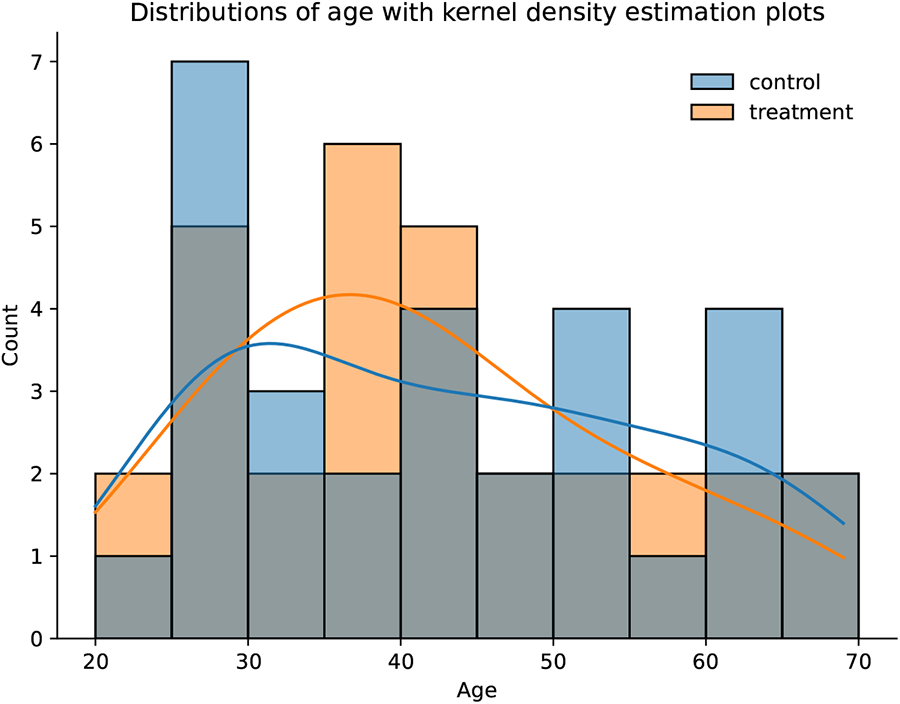
The distributions of age for the control and treatment groups with kernel density estimation curves: 30 people in each group, 60 people in total.

Figure 2 presents the distributions of PANSS subcategories and the total PANSS scores for the evaluated patients. Mean values with corresponding standard deviations for PANSS positive, negative, general, and total score are 16.37 *±* 6.03, 19.80 *±* 8.34, 35.30 *±* 9.71 and 71.47 *±* 20.21, respectively. Comparison of these statistics with Table 1 from [Benjamin et al., 2021] implicates that the treatment group in our study has more severe symptoms of psychosis. Patients were excluded from this study if they met at least one of the following criteria: heart conduction disorder, epilepsy, past cardiac surgery and/or neurosurgery, past head injury with loss of consciousness, chronic use of beta-blockers, cognitive disorders precluding cooperation or showing a lack of cooperation as well as having an allergic reaction to the wearable sensor. All patients had an ongoing disease history and were diagnosed with schizophrenia or BD by a qualified psychiatrist in accordance with the ICD-11 guidelines. Members of the control group had no diagnosed, ongoing mental disorder and were not taking antidepressants or heart-rate-affecting beta-blockers.

**Figure 2:**
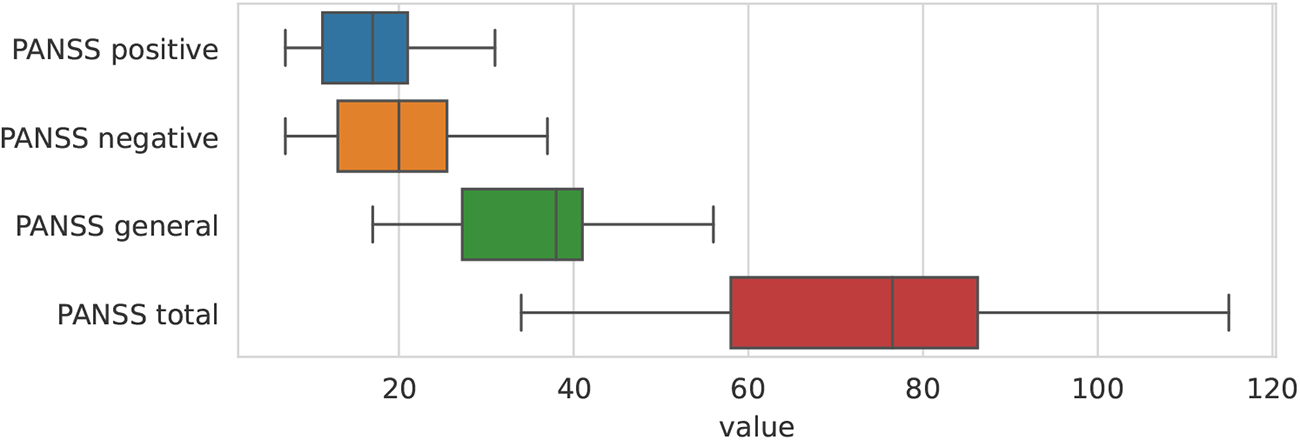
The distributions of values of the considered PANSS subcategories (assessing the severity of positive, negative and general symptoms) and the PANSS total score (the sum of all subcategories) based on data from 30 patients.

All participants were equipped with a Polar H10 chest strap for about 1.5–2 hours and instructed to carry on their daily routines with additional encouragement to undertake physical activity to the extent of taking a walk. Due to the patients being in the medical ward, they received instruction to walk in the corridors at the beginning of the experiment, after 30 minutes and 1 hour. The subsequent walks were to be slightly longer (about 3, 5, and 7 minutes, respectively). Some participants were not able to wear the device for such a long period and their measurements were shorter (not less than 70 minutes). Additionally, at the start, participants were requested to rest in a sitting position for 5 minutes. However, strict following of these instructions was not enforced during the experiment, both in the treatment and the control group.

After ECG data collection, patients underwent psychiatric evaluation using a Polish-validated PANSS questionnaire [Rzewuska, 2002]. It is a scale and structured interview questionnaire used to assess the severity of positive and negative symptoms in patients with psychotic disorders, mainly schizophrenia, commonly used for evaluation in research purposes [Opler et al., 2017]. The scale consists of 30 items describing the patient’s symptoms, divided into three categories: positive symptoms (7 items), negative symptoms (7 items), and general psychopathological symptoms (16 items). In each item, the examiner can give the symptom a value from 1 to 7 based on its described severity, where one indicates the absence of symptoms and seven indicates their extreme severity. Within this scale, a patient can receive a score ranging from 30 to 210 points, and the assessment takes approximately 45 minutes [Andreasen et al., 1995].

### 2.1 Estimation of patient HRV from ECG data

HRV values were calculated from N-N intervals using the Root Mean Square of the Successive Differences (RMSSD) approach. RMMSD is a time domain method corresponding to the high-frequency part of the HRV spectrum that ranges from 0.15 to 0.4 Hz. A single HRV value can be derived through the following equation:

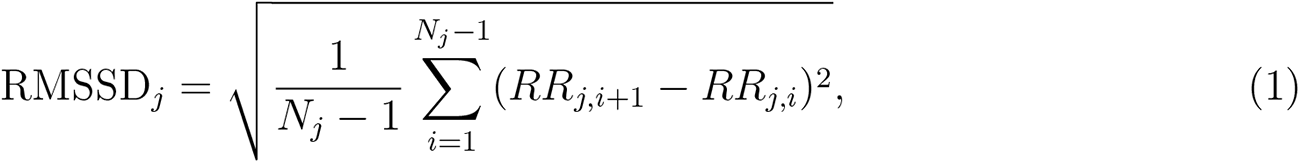

where RMSSD*_j_* is the *j*-th HRV value, *j ∈ {*1*, …, M }*, and *M* is the number of the time windows in the whole series, *RR_j,i_* is the *i*-th N-N interval (corrected R-R interval) value in the *j*-th time window, *RR_j,i_*_+1_ *− RR_j,i_* represents the difference between two consecutive N-N intervals, and *N_j_* is the number of N-N intervals in the *j*-th time window.

### 2.2 Estimation of patient mobility from accelerometer data

Accelerometer data contains a three-axis measurement of proper acceleration (i.e. acceleration of a body in its own instantaneous rest frame). As such, it contains both gravity and mobility components. To isolate the latter, the momentary gravity component was estimated using a low pass filter and subtracted from the registered acceleration data. The absolute value of a mobility acceleration component was then integrated within a time window corresponding to the R-R interval to get a numerical mobility estimate.

The accelerometer data was sampled at 100 Hz within *±*4g range. For momentary gravity component estimation, a Gaussian filter with *σ* = 10^3^ was used (corresponding to a window of about one minute of data). Since the accelerometer sampling rate is higher than HRV values, downsampling of its data was performed.

### 2.3 Data collection

A Polar H10 chest strap was used to collect R-R intervals and accelerometer data. The quality of its R-R interval measurements during different activities (e.g., sitting, walking, training, etc.) was analyzed by [Gilgen-Ammann et al., 2019] and assessed to match 99.6% of the true R-R intervals. The evaluation took into account the wrong detection of R-wave peaks or their missing detections. The results indicate this wearable device may be reliable enough to measure HRV in patients during different activities.

We used Polar Sensor Logger [Happonen] for the collection of raw data: ECG, R-R intervals, accelerometer and heart rate values. The implementation to reproduce all experiments and results is available under an open license Książek et al. [2023a] and the dataset is publicly available in the Zenodo repository Książek et al. [2023b].

### 2.4 Data correction

The recorded R-R intervals contain artifacts related to ectopic heartbeats and sensor issues, e.g. loosening the device on the chest or disconnection from the smartphone. The presence of artifacts may introduce bias in results, including abnormally high standard deviation of HRV values [Thuraisingham, 2006, Peltola, 2012]. Therefore, we prepared a multi-stage data correction according to the suggestions from [Peltola, 2012]. We combined automatic anomaly detection using a wavelet transform [Keenan, 2008] with manual artifact removal. Manually deleted measurements include values obtained with a delay or mistaken timeframe due to equipment error. We also removed the first and last 45 seconds of the recording since installation and detaching the device, respectively. Moreover, we excluded 15 seconds of measurements after at least a 30-second gap caused by connection problems. Then, Daubechies wavelet transform (DWT) was applied with a filter length of 5 and all heartbeats in the 5-second neighborhood of the measurements indicated as anomalous by DWT were removed. We also removed the first R-R value after every gap longer than 2s. Finally, after the visual inspection, the remaining suspicious heartbeats were removed manually.

### 2.5 Interpolation

After data correction, in selected settings, data was interpolated with cubic splines. The interpolation is a recommended stage in the HRV calculation process [Quintana et al., 2016] and is strictly necessary when frequency analysis is performed. However, for long gaps, the interpolated data may distort HRV values. Therefore we compared both interpolated and non-interpolated time series.

### 2.6 Data windowing

A rolling window was used to divide data into partially overlapping subseries. The method has two parameters: window length and a time interval between consecutive subseries (in minutes). Neighboring windows may contain common elements, e.g. for window length *M*, time interval *N*, and the *i*–th subseries if only *N < M*, the (*i* + 1)–th window will contain data from the last *M − N* minutes of the *i*–th window. We do not consider time window setups for which *N > M* because a part of the data would be omitted. A median value was selected as a timestamp for a time window due to the presence of data gaps (caused, e.g. by device disconnection), which would make a mean timestamp value unrepresentative.

### 2.7 Evaluation methods

Pearson correlation coefficient and corresponding p-values between the mean HRV values in consecutive time windows and the PANSS total score and its subcategories (positive, negative, and general symptoms) were computed. Since many statistical tests were performed for the same HRV series, the Bonferroni correction has been applied. The adjusted p-value threshold equals 0.125 which corresponds to the typical significance level *α* = 0.05.

## 3 Results

### 3.1 Comparison between the tested groups

To verify the existence of a statistically significant difference in mean HRV values between the treatment and the control group, a Mann-Whitney U (MWU) test for two groups of samples was applied. MWU was used instead of the t-test because, according to the results of the Shapiro-Wilk test, the assumption about the normality distribution of data samples within the groups was not met. A one-tailed test with the adjusted significance level *α* = 0.025 was used to evaluate whether mean HRV values for patients are significantly lower than those for people from the control group. The calculated statistic *U* = 83 with p-value *≈* 3.01 *·* 10*^−^*^8^ indicates that the null hypothesis about equality of means in two groups can be rejected in favor of the alternative hypothesis that HRV values of the treatment group are lower than those of the control group. The equality of variance between the two compared groups was also evaluated. The results of Levene’s test suggest that the two considered variances are significantly different (the test statistic *W ≈* 11.62, p-value *≈* 0.001). Figure 3 presents a detailed comparison of HRV distributions for both groups. The median HRV value of the treatment group was equal to *≈* 8.73 (with standard deviation *≈* 3.87) while the median HRV value of the control group was *≈* 18.72 (with standard deviation *≈* 7.54). Results indicate that both patient’s mean HRV value and its variance are lower than in the control group.

**Figure 3:**
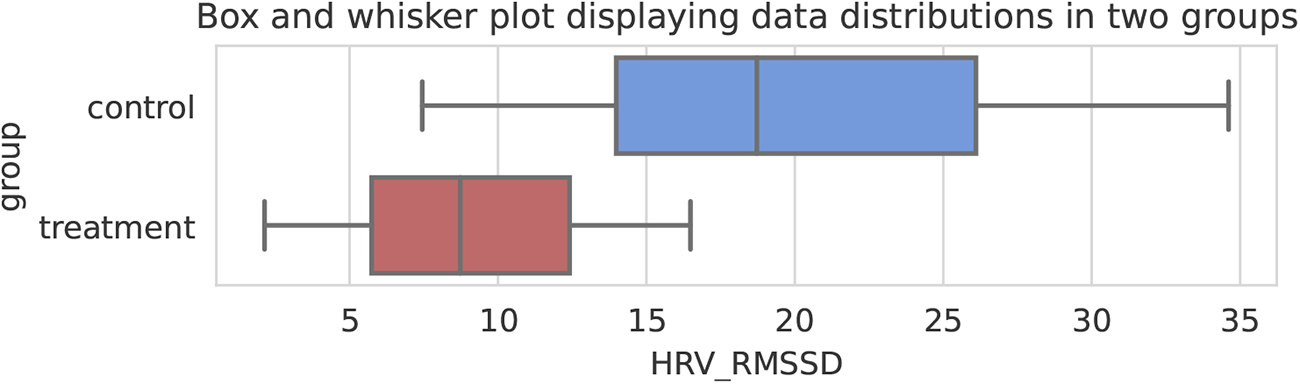
Box and whisker plot presenting the distributions of mean HRV values for the treatment and the control groups compared in the study.

### 3.2 Relationships between HRV and PANSS test scores

Figure 4 presents relationships between mean HRV and PANSS test scores for 30 patients in terms of Pearson’s r with 95% confidence intervals. In this case, a rolling window of 15 minutes with 1-minute time intervals between consecutive windows was used and data was not interpolated. The relationships were measured separately for each PANSS subcategory.

HRV and PANSS general values are statistically significantly correlated, even after the Bonferroni correction (*r* = *−*0.453, p-value = 0.012). In the remaining subscales, the highest effect size was noted between HRV and PANSS positive/PANSS total scores, i.e. *−*0.317 and *−*0.297, respectively. However, these dependencies were not considered statistically significant. The result for PANSS negative scale suggests no relationship between negative symptoms in schizophrenia and HRV values.

Despite the fact that a limited number of samples were available (30 patients), our observations are consistent with the results presented in Table 2 in [Benjamin et al., 2021]. The authors achieved the highest effect size comparing HRV with PANSS general (*r* = *−*0.32) and the lowest one for PANSS negative scale (*r* = *−*0.1). Only the correlations for PANSS general and PANSS total were statistically significant (p-value was equal to 0.003 and 0.03, respectively), while the highest p-value was related to PANSS negative (p=0.37).

### 3.3 Sensitivity analysis

Figure 5 presents the impact of hyperparameters on the relationship between the mean patient’s HRV and the results of the PANSS general score measured by the Pearson correlation coefficient. HRV was calculated in time windows (*window size*) of range *{* 1, 2, 3, 5, 10, 15, 20*}* minutes while the time interval between consecutive windows (*step*) had a length of *{*0.25, 0.5, 0.75, 1, 2, 5, 10*}* minutes. If in a given pair of hyperparameters *step* was greater than *window size*, then this setup was excluded from calculations because some portions of data would be omitted. Bold font represents cases for which the p-value was lower than 0.0125. For cases where data interpolation has been omitted, the individual p-values ranged from 0.0107 to 0.0213, while with data interpolation, they varied from 0.0010 to 0.0170. We observed a tendency that for greater time windows and larger time intervals, the effect sizes were increased. The highest effect size was noted for 15-minute time windows and 10-minute steps between them. It reached a value of *−*0.459 without data interpolation and *−*0.465 in the opposite case. The results suggest that short HRV windows may be prone to local variations. This experiment is intended as a discussion of a general tendency for different hyperparameters and should be considered auxiliary because, after the application of the Bonferroni correction for multiple comparisons, the statistical significance is lost.

**Figure 4:**
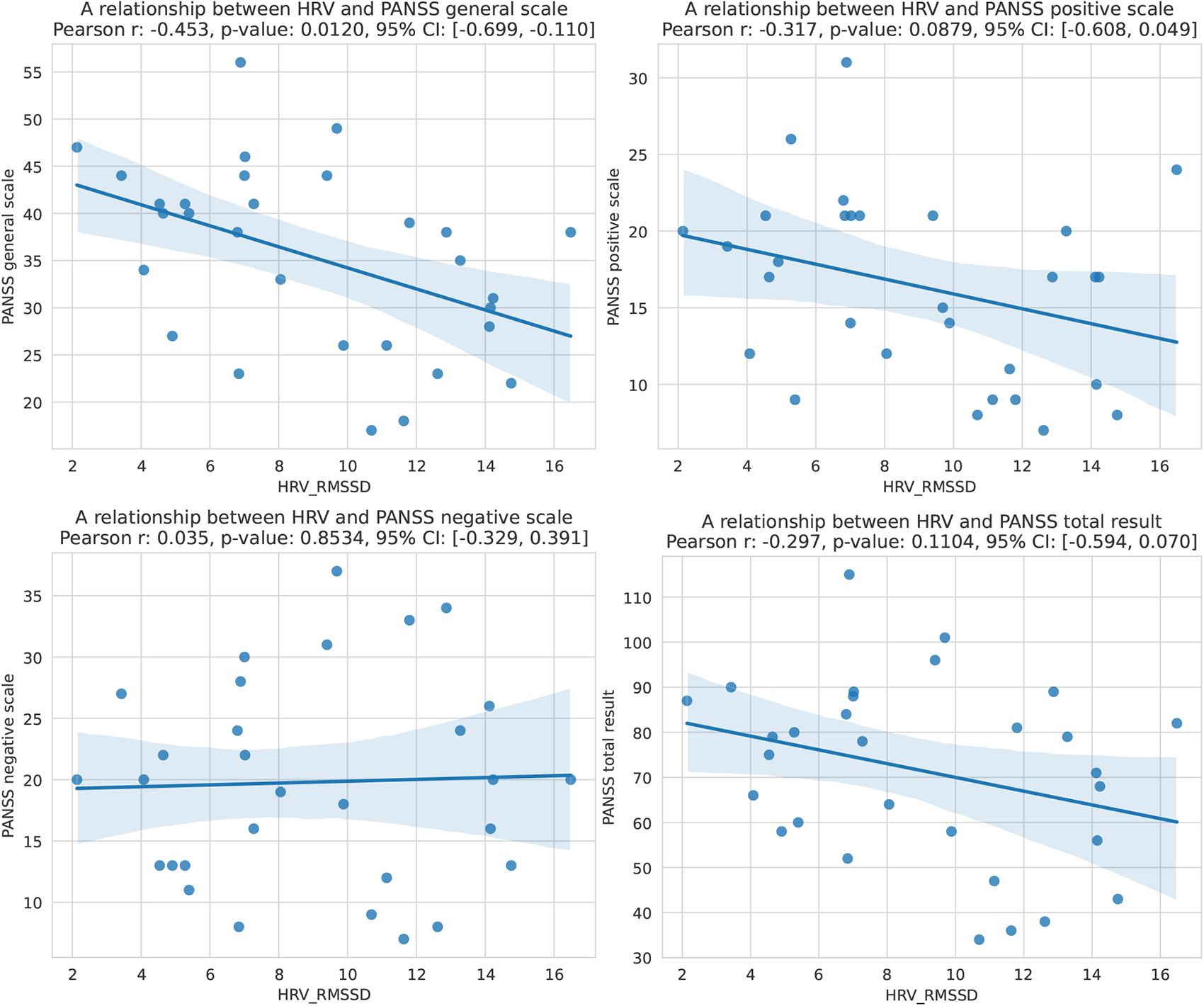
Relationships between mean HRV values (in terms of RMSSD) and PANSS in general, positive, negative scale or PANSS total result for 30 patients. Consecutive scores present regression lines with 95% confidence intervals. A single point corresponds to one person from the treatment group. Calculations were made for 15-minute rolling time windows with 1-minute intervals between them, without data interpolation.

### 3.4 Mobility analysis

The literature suggests that HRV negatively correlates with heart rate [Kazmi et al., 2016, Yiiong et al., 2019]. To verify this, an analysis based on accelerometer data collected together with R-R intervals was performed for both groups. Figure 6 presents an example of HRV and mobility coefficient values for one of the persons from the control group. In this case, HRV is inversely proportional to the mobility values. However, an interesting phenomenon was observed. Figure 7 presents a scatter plot illustrating a relationship between mean HRV and Pearson correlation coefficient values calculated for every person. Each point corresponds to a person from the treatment (red) or the control group (blue). For almost all (28 out of 30) members of the control group, the correlation values are negative. This is true for only 21 out of 30 patients in the treatment group. Patients have significantly lower HRV than control participants. Furthermore, all members of the treatment group with positive correlation have a mean HRV value below 10, which is clearly visible in the bottom-right corner of the plot. Moreover, among the participants with a positive correlation, the highest HRV values belong to the members of the control group. This indicates that HRV and mobility together may be promising markers in disease diagnosis. However, further studies with more participants are necessary to draw more detailed conclusions.

**Figure 5:**
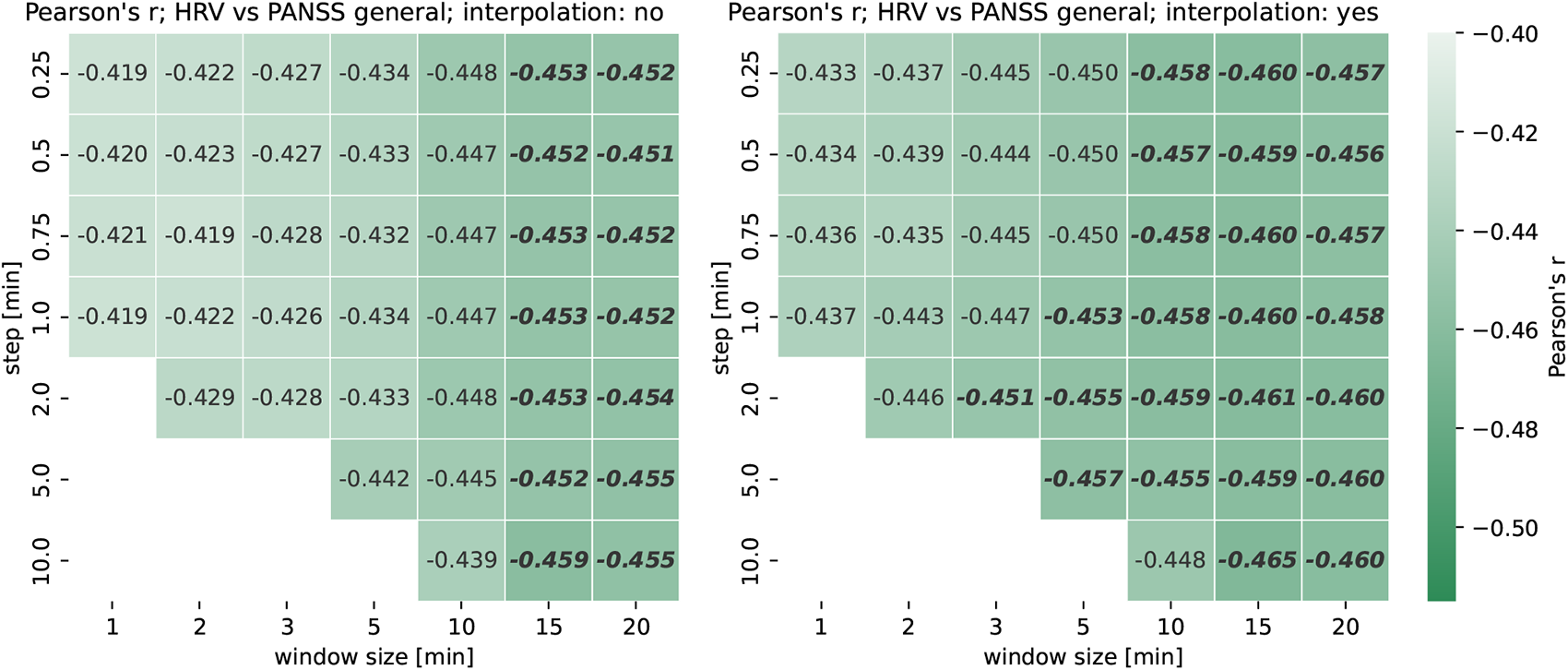
A dependency between HRV and PANSS general for different configurations of HRV calculation, in terms of Pearson’s r. Bold font refers to the results having p-values lower than 0.0125. *Step* derives width between consecutive time windows while *window size* denotes the time range of a particular rolling window. Cases in which *step* was greater than *window size* were neglected.

**Figure 6:**
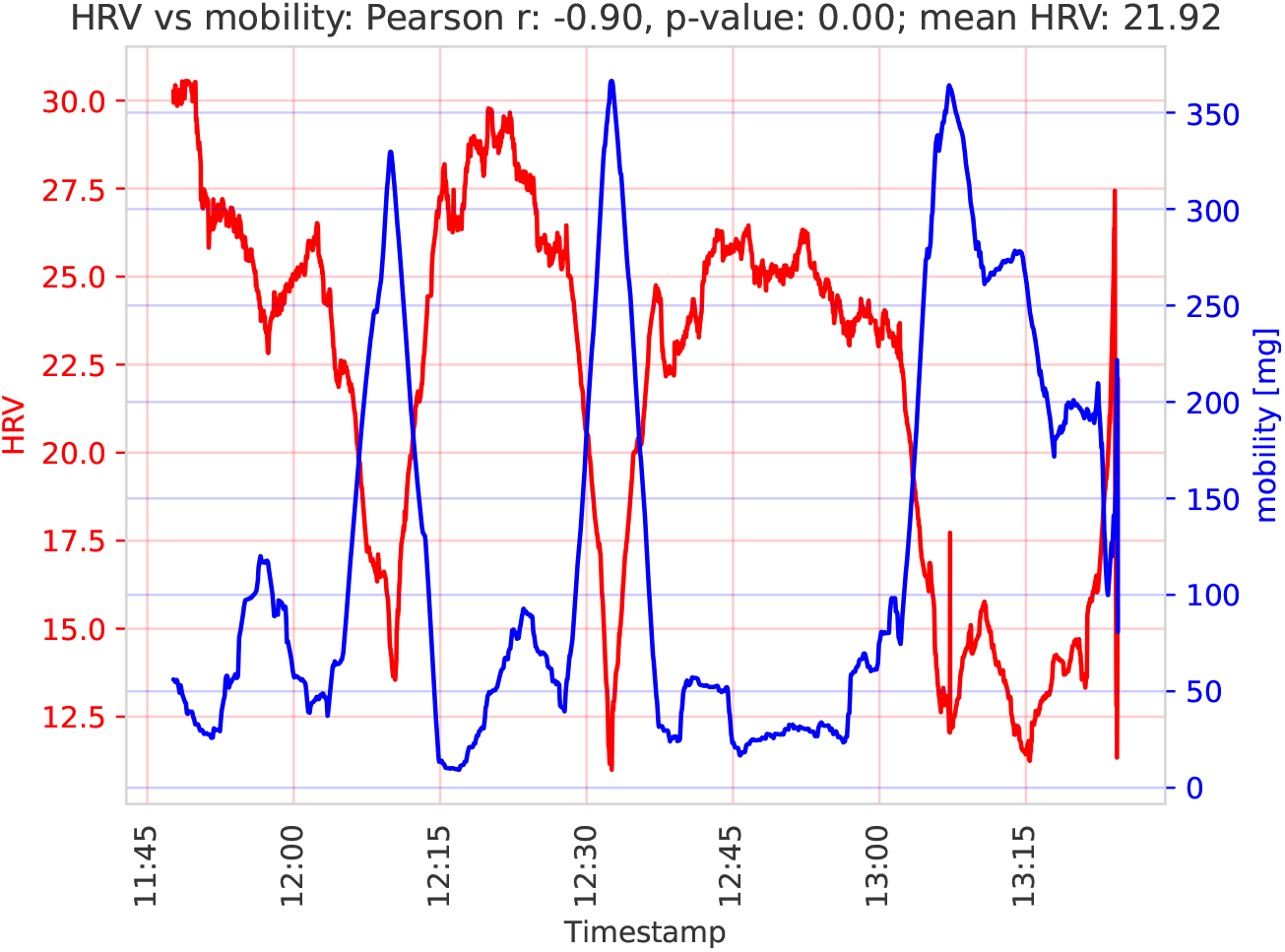
Values of HRV and mobility coefficients in subsequent time steps for a selected person from the control group.

**Figure 7:**
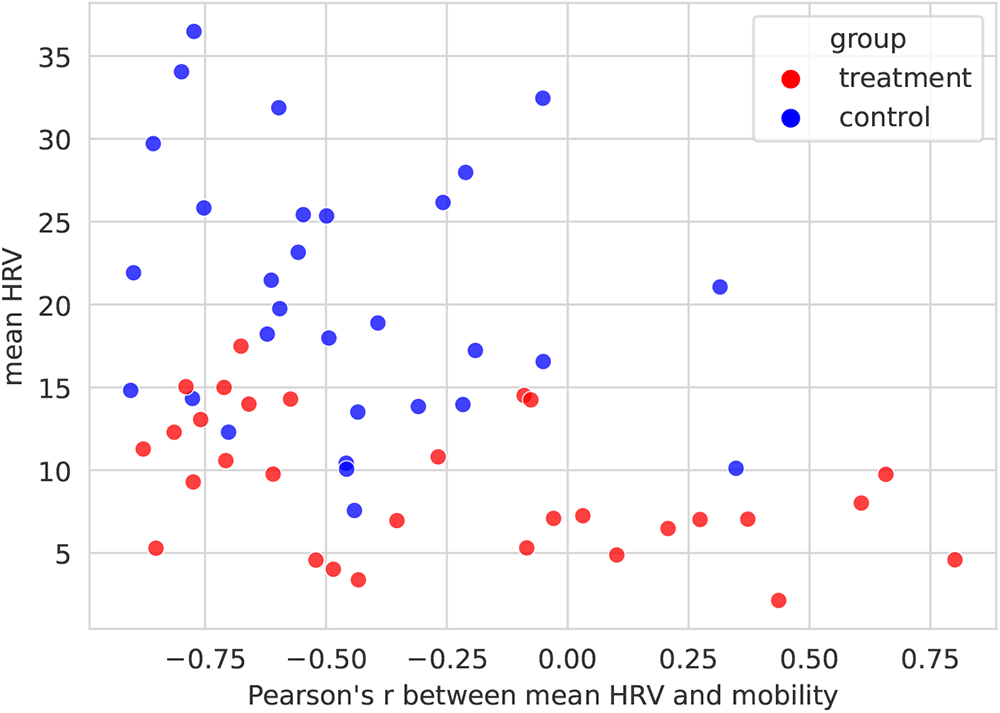
A relationship between mean HRV and Pearson’s r measuring the association between HRV and mobility of a given person calculated based on accelerometer data.

## 4 Discussion

### 4.1 General remarks

Our findings are consistent with [Benjamin et al., 2021], where the authors noted that the HRV values for the schizophrenia patients were lower than for the control group. This was more difficult to observe for bipolar disorder patients. Due to the limited number of samples, we did not perform separate comparisons for the two groups of patients. However, in the paper [Benjamin et al., 2021], the correlation coefficient was statistically significant between HRV and the PANSS test in terms of the total score and general subscale while in our study, the relationship was significant only in the second case. Our results are also in line with [Clamor et al., 2016] where a meta-analysis was performed, indicating that patients with schizophrenia (a total of 1016 individuals) had lower RMSSD values than people from the control groups (1469 persons). However, some results indicated a significant decrease in HRV only for patients with bipolar disorder and not in schizophrenia patients [Henry et al., 2010]. In general, our results are consistent with the literature that suggests a relationship between psychosis severity and HRV as a marker of significantly impaired ANS functioning [Stogios et al., 2021].

### 4.2 Mobility measurement as a part of HRV assessment

Different approaches exist as to what time frame to select for HRV assessment [Task Force of the European Society of Cardiology and the North American Society of Pacing and Electrophysiology, 1996], with propositions ranging from single-digit minutes to double-digit hours. With the latter range, a variation in patient mobility (and thus heart rate) can be safely assumed. In a general population, a negative correlation between heart rate variability and heart rate has been observed. In [Kazmi et al., 2016] this effect has been noted both in humans (across normal sinus rhythm and congestive heart failure) and in animals. A similar relationship has been noted while investigating biometric parameters of exercise movements [Yiiong et al., 2019] – a negative correlation of HRV and motion parameters has been observed.

Our results support this relationship for the control participants (see Figure 7, control group, a negative correlation of HRV and mobility). However, we observed an inverse or no relationship for a subset of our treatment group. While this paints a complex picture of the relationship between mobility and HRV for diagnosed patients, the observable differences mark a promising avenue of investigation. Motion is easy to capture with ubiquitous devices, and most patients are able to perform at least basic mobility tasks. There is potential for using this relationship for diagnosis. Research by [Reinertsen et al., 2017] and [Osipov et al., 2015] show that a combination of heart rate and activity features outperforms using either alone in schizophrenia classifier performance.

An important aspect of HRV and mobility relationship is the time assigned for measurements. When observed in 24h intervals, this dependency may be much less pronounced – in [Hautala et al., 2010], a very limited correlation has been observed over 24h intervals, showing a tendency similar to our treatment group. However, more significant relationships between HRV and physical activity were noted for short-term HRV calculations. They are in agreement with our findings based on 1.5-2 hours of measurements for the control group. The authors have limited the study to healthy persons while we observed a different tendency in patients. Interestingly, according to results in [Bengtsson et al., 2021], mobility could not be used as a predictor for the difference in HRV between patients with depression, schizophrenia, and the control group, even while ongoing physical activity was found to have a significant relationship with HRV. It is worth investigating whether such apparent contradiction with our results can be traced to individual (personal) differences, similar to those seen in our Figure 7.

To our knowledge, there are no studies that analyze the relationship between accelerometry values and HRV in schizophrenia or BD in a similar manner to ours. Available research that takes into account both heart rate parameters and accelerometry for the assessment of schizophrenia or BD tend to: use heart rate instead of HRV as a general parameter [Osipov et al., 2015, Reinertsen et al., 2017, Carr et al., 2018], compare a short 5 min HRV reading with general activity [Freyberg et al., 2020], use accelerometry only for artifact detection [Stautland et al., 2022] or collect the data simultaneously but analyze them separately [Faurholt-Jepsen et al., 2017, Cella et al., 2018]. An experimental protocol that could use an accelerometer and HRV data in a similar way to ours was also registered, although the results are yet to be published [Cotes et al., 2022].

### 4.3 Effect of quetiapine on HRV

In the study [Hattori et al., 2018], the authors considered the impact of different antipsychotics on autonomic nervous system functioning. They compared patients taking risperidone, olanzapine, quetiapine and aripiprazole. Their analysis indicated that quetiapine might reduce low and high-frequency components of HRV more significantly than the remaining medicines. Therefore, we prepared an additional study that excludes seven patients taking quetiapine from our 30-person treatment group. The experiment results are presented in Figure 8. The strength of the negative relationship between mean HRV and PANSS in general scale increased from 0.453 to 0.475. However, it was related to the growth of the p-value. It is worth noting that none of the patients taking quetiapine achieved a mean HRV higher than 12. It is also necessary to emphasize that our findings are very limited due to a small sample and an extensive study with more patients should be performed.

## 5 Conclusions

At this stage, HRV analyzed according to the RMSSD approach cannot be used to diagnose and treat schizophrenia or BD and function as a substitute or add-on to a psychiatric interview. However, there is a possibility that with the combination of such factors as more complex HRV analysis methods, incorporation of accelerometers for activity analysis, and consideration of the effect of psychotropic medications, HRV along with mobility coefficient may become a sufficient biomarker for diagnosis and treatment of psychotic disorders such as schizophrenia or BD. The correlation between HRV and some PANSS scores and dependencies found between the mobility coefficient and HRV suggest that this approach may be auspicious. However, it needs larger data samples and further experiments.

**Figure 8:**
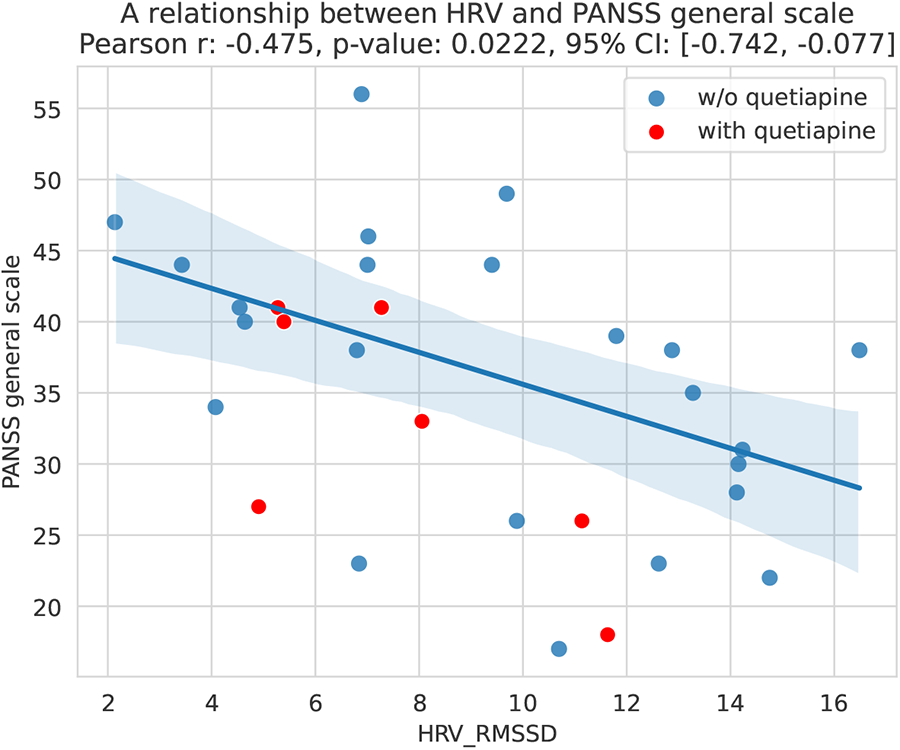
A relationship between mean HRV values and PANSS in general scale for 23 patients not taking quetiapine (blue points). The plot presents a regression line with a 95% confidence interval. For reference, scores of 7 patients taking quetiapine (red points) are presented but were not included in this study. Compare with Figure 4 where all patients were considered. The experiment used 15-minute rolling time windows with 1-minute intervals between them, without data interpolation.

## Data availability

The implementation to reproduce all experiments and results is available under an open license (https://github.com/iitis/Polar-HRV-data-analysis) and the dataset is available in the Zenodo repository (https://www.doi.org/10.5281/zenodo.8171266).

